# Simulating genetic risk scores from summary statistics with an application to type 1 diabetes

**DOI:** 10.1101/2024.05.17.24307282

**Authors:** Steven Squires, Michael N. Weedon, Richard A. Oram

## Abstract

**Motivation:** Genetic risk scores (GRS) summarise genetic data into a single number and allow for discrimination between cases and controls. Many applications of GRSs would benefit from comparisons with multiple datasets to assess quality of the GRS across different groups. However, genetic data is often unavailable. If summary statistics of the genetic data could be used to simulate GRSs more comparisons could be made, potentially leading to improved research.

**Results:** We present a methodology that utilises only summary statistics of genetic data to simulate GRSs with an example of a type 1 diabetes (T1D) GRS. An example on European populations of the mean T1D GRS for real and simulated data are 10.31 (10.12-10.48) and 10.38 (10.24-10.53) respectively. An example of a case-control set for T1D has a area under the receiver operating characteristic curve of 0.917 (0.903-0.93) for real data and 0.914 (0.898-0.929) for simulated data.

**Availability:** The code is available at https://github.com/stevensquires/simulating_genetic_risk_scores.

## Introduction

A genetic risk score (GRS) provides a summary of variation in the genome that are related to some trait of interest [4]. Generally these are produced by application of genome wide association studies (GWASs) [11] that search through genetic variants looking for parts of the genome that are associated with that trait and provides both the statistical significance and the strength of the association.

A GRS is produced by taking the genetic variants that show a statistically significant association with a trait. The associated variant weights (strength of the association) are summed with the number of alleles of that variant (out of the two chromosomes). There are also more complex GRSs, for example including interaction terms between variants which turn the standard linear GRS into a more complicated non-linear GRS [6]. A useful GRS will provide good discrimination between the groups of samples with the trait from those without.

The primary motivation for this work is to make the generation of GRSs more accessible for research. A common research problem might be about how a specific GRS works on some particular dataset, or on multiple datasets. As the purpose of a GRS is the capacity to discriminate between samples with and without a trait, making multiple comparisons can be essential. For example, if an aim is to test an existing or new GRS it is likely that it will require multiple datasets to provide a more confident judgement about its performance. Datasets of interest might include: different population ancestry, variants of a disease, geographic locations, or socio-economic status.

There are two major issues with generation of GRSs in multiple datasets: one is access to the data and the second is the expertise required to correctly generate the GRS. The first issue can be difficult because of, for example, ethical [10], legal [1] or computational [7] reasons. The second issue is that genomic data tends to be held in specific file formats and requires particular software to be effectively run, requiring people with specific skill-sets. This is a barrier for groups without this expertise to utilise GRSs.

Our aim is to provide a GRS simulation method from summary statistics. Ethical and legal requirements can be significantly reduced as only a small amount of data about the dataset needs to be released to enable the production of simulated data. A range of scores on multiple datasets can be produced to enable much wider comparisons. Significantly less specific technical skills are required as the summary statistics are all that is needed to generate the GRSs along with the provided Python code. If summary statistics could be collected and stored in one place then it would be possible to automatically produce datasets of GRSs from just those summary statistics.

We provide a methodology to take easily accessible summary statistics and generate simulated complex GRSs without the need for access to the original genotype array data. Our method requires: single nucleotide polymorphism (SNP) frequencies, correlations between SNPs, and any SNPs that deviate from Hardy-Weinberg equilibrium (HWE). Both simulated SNP arrays (for the SNPs in the GRS) and the GRS are generated from these summary statistics. We demonstrate that our simulated model produces similar results to real data for the final GRS, relevant sub-parts of the GRS, and distributions of the SNP values.

## Methodology

### Overview

Our simulation approach comprises three overall steps: 1) collection of necessary summary statistics; 2) generation of the simulated SNP array; 3) production of the final GRS. We briefly describe the overall method and show the steps in Algorithm 1 before going into detail subsequently.

#### Algorithm 1 Method overview

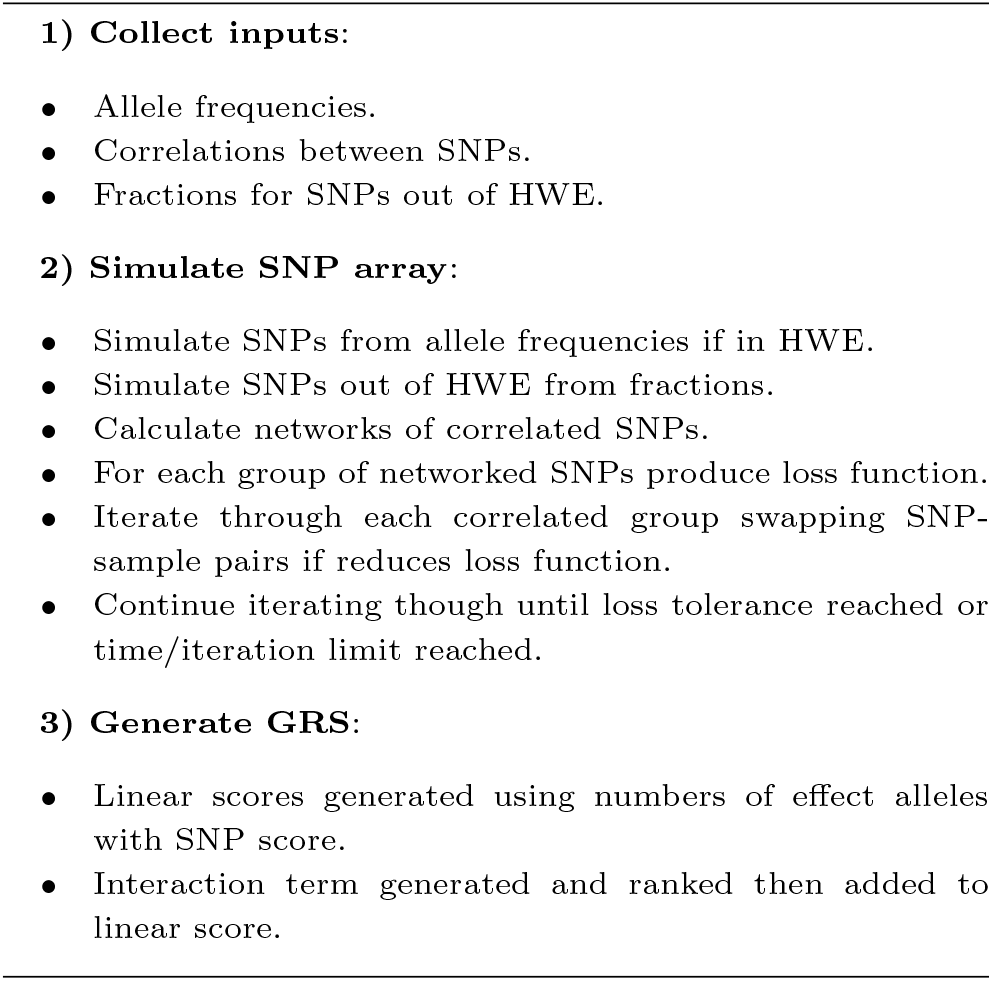

There are three summary statistics required: the allele frequencies of each of the SNPs; the correlations between all the SNPs; the proportions of homozygous and heterozygous alleles if any of the SNPs deviate from Hardy-Weinberg equilibrium (HWE). These can either be accessed by application to a server producing these summary statistics, or by generation from the original SNP arrays.

The aim of simulating SNP arrays is to produce individual samples that are indistinguishable from real data. Our approach is to simulate SNP arrays that have both the same allele frequencies and the same correlations between SNPs as the real data.

We perform this in several stages. First, *N* samples are generated from the allele frequencies with any SNPs that deviate from Hardy-Weinberg equilibrium generated separately based on the proportions of homozygous and heterozygous alleles. Secondly, the correlated SNPs are collected into *G* groups of correlated SNPs. Within each group the correlations between the SNPs are calculated and a loss function is defined. Pairs of each correlated SNP in turn (across two samples) are compared and switched if they reduce the loss function. This approach is continued until the loss function falls below a chosen tolerance. The same procedure is applied for all *G* groups which results in the final simulated SNP array.

As an example of GRS production we use a type 1 diabetes (T1D) GRS [8] that includes both a linear term and a term related to interacting SNPs. These two parts are generated separately from the simulated SNP array and combined together. Our simulated GRS model would work on any linear model (the interaction terms are excluded) or any GRS with both a linear and interaction part that is similar to this T1D GRS.

### Datasets and GRS

To develop our method and generate results we need the relevant summary statistics. To test that the simulation approach works effectively we need to also have real SNP array data so we can make direct comparisons.

We use two publicly available datasets: the 1000G version 3 data [2] and UK Biobank [9] SNP array data. The 1000G dataset consists of 2,503 samples from five super-populations: Europeans (EUR), Africans (AFR), Americans (AMR), east Asians (EAS), and south Asians (SAS). These five super-populations are made up of a total of 26 populations with 4 to 7 populations making up each super-population. We consider five subsets of the UKBB data. Four of these are defined by applying principal component analysis (PCA) to the SNP arrays and defining the subsets by similar locations in the first two principal components to four of the 1000G data (EUR, AFR, EAS, SAS). The fifth subset is European samples with T1D.

As a demonstration of the effectiveness of the method we consider a T1D GRS [8] which consists of 67-SNPs. The GRS involves both a set of linear weights on the number of effect alleles and an additional pair-wise interaction term based upon which of 18 pairs of 14 SNPs each sample has. Importantly the GRS contains multiple SNPs from part of the genome where there are known to be substantial correlations between the SNPs (within the human leukocyte antigens, HLA, region). Therefore, it is important when generating simulated data to fully include these correlations between SNPs to ensure we are generating a set of SNP array data which is similar to the real data.

Due to missing SNP data in the 1000G dataset we exclude two SNPs leaving 65 in the GRS. For the UKBB data we use all 67 SNPs.

### Collection of Summary Statistics

We collect summary statistics from two datasets: the 1000G and UK Biobank. For 1000G we use the NIH LDlink suite of applications [5] to extract variant frequencies and linkage disequilibrium between SNPs. We also use the 1000G SNP array data to extract the SNPs that deviate from Hardy-Weinberg equilibrium.

For UK Biobank we use imputed array data extracting the required SNPs and taking the frequencies and correlations from the arrays. We use similar positions in 2-dimensional PCA space as the 1000G super-populations.

### Generation of Simulated SNP Arrays

We apply two constraints to the SNP arrays, the first is that the frequencies of the alleles need to match the summary statistics from the real data. Secondly, the correlations between the SNPs need to also match the known correlations. There may be a third constraint if any of the SNPs are out of Hardy-Weinberg equilibrium, for example due to population stratification. If that is the case then we need to impose an additional constraint to set the correct proportions of homozygous and heterozygous alleles for these SNPs.

The key aspect of our method is that the allele frequencies (and any deviations in homozygosity and heterozygosity) remain constant under permutations of the SNPs between samples. So if we set up a matrix of sample and SNP values that has the correct SNP frequencies (and homozygosity deviations) then if we permute individual SNP-samples (i.e. switch over SNP values between samples) in the matrix we do not alter the frequencies but can change the correlations between SNPs.

We define the simulated SNP array as a matrix **X** ∈ *R*^*N*×*p*^ where *p* is the number of SNPs in the GRS and *N* is the number of desired simulated samples. We consider each SNP in turn and generate the simulated data in one of two ways. If the SNP holds to HWE then we generate it from the frequency by the generation of two binary *N* -dimensional vectors (one for each chromosome) with probabilities set by the allele frequency, the SNP values are then the sum of the two. If the SNP deviates then we have the fractions of the homozygous and heterozygous values and we generate fractions of 0s, 1s and 2s as required.

On generation of the SNPs we have a matrix of size *N* × *p* of 0, 1, 2s but with no correlation between the SNPs. The SNPs with no expected correlation are left alone. For the rest of the SNPs we group them into those that are correlated together. In Figure 1 we show an example of correlated SNP groups. The nodes (labelled A-G) represent SNPs and the lines represent those SNPs which are correlated together. So {*A, B, C*} are correlated together but neither *A* or *B* are correlated with D or E. E is only correlated with D. D is correlated with both C and E. F and G are correlated together but neither is correlated with any of the other SNPs. As the pair of groups {*A* − *E*} and {*F* − *G*} are not correlated together we can optimise them separately.

**Fig. 1.**
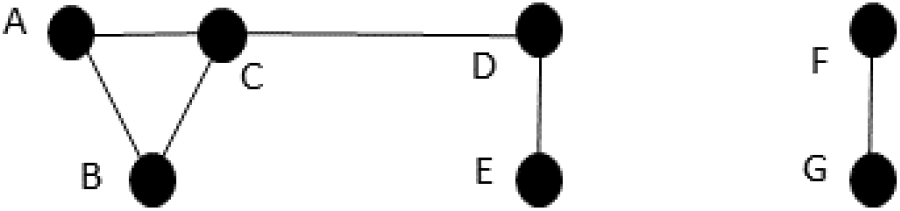
Examples of correlated SNPs. The two groups A-E and F-G are not correlated together. Within group A-E: A, B and C are all correlated together; C is additionally correlated with D; D is correlated with C and E; E is just correlated with D. F and G are correlated together and neither is correlated with any other SNPs.

To find the correlated groups we define a correlation threshold between SNPs above which the SNPs are defined as correlated (with number of SNPs, *p*_*C*_) and below which are defined as uncorrelated. We build an adjacency matrix for all *p*_*C*_ SNPs and use the Python network analysis package NetworkX [3] to extract the set of groups of correlated SNPs, *G* = {*g*_1_, …*g*_*m*_}. Each *g*_*i*_ group of SNPs are connected by correlations between then, for example the SNPs {*F, G*} or {*A, B, C, D, E*} from Figure 1.

We now have a set of groups of SNPs. We perform the same procedure separately for each, *g*_*i*_, group. We set up a loss function which is defined by

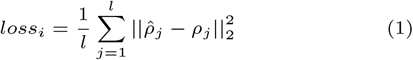

where the sum is over the correlated pairs of SNPs (from 1 to *l*), 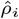 is the correlation between the pair of simulated SNPs and *ρ*_*i*_ is the real correlation between the SNPs. For example the loss function for the left side set A-E of SNPs in Figure 1 would be given by

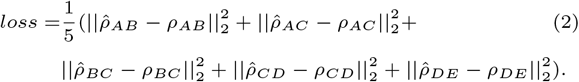

For a group of correlated SNPs, *g*_*i*_, the procedure is as follows. We consider all possible SNP pairs in *g*_*i*_ and randomise the order we consider the SNPs. We then take a random permutation of the N samples and compare the neighbouring (randomised pairs) of samples, for that pair of SNPs. We swap the SNP values across the pair of samples and check if the loss function has reduced, if it has we make the change permanent, if not then we revert to the previous state. The algorithm then moves onto the next pair of samples and performs the same procedure. If at any point the loss falls below the defined tolerance the iterations are stopped and the SNP values are set. Once all the pairs of randomised samples have been iterated through (and if the loss is still above the tolerance level) the same procedure is applied to the next pair of SNPs (if there are more than two SNPs in the group). Once the procedure has been applied to all pairs of SNPs, the order of SNPs is randomised so different pairs of SNPs are compared in order. A new random permutation of samples is set up and the iteration through pairs of samples continues. There is also a set iteration or time limit that can be imposed.

Part of this process is shown, as a toy example, in Figure 2. There are two correlated SNPs (the two columns) with four samples (the four rows). The algorithm first swaps *x*_11_ and *x*_21_ (step 1). It then calculates the new loss function and it has not fallen therefore in step 2 the pair of SNPs are returned to the original positions. The algorithm then (step 3) swaps *x*_11_ and *x*_31_ and checks the loss function. This time the loss function is lower so it leaves the SNP array in the new permutation.

**Fig. 2.**
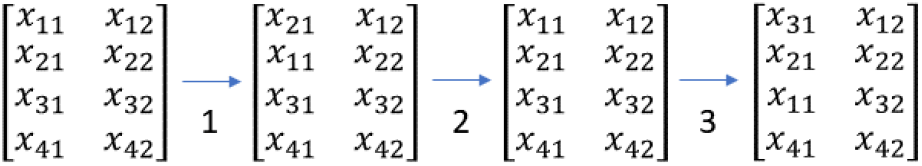
A toy example showing the algorithm being applied to two correlated SNPs with four samples. In step 1 *x*_11_ and *x*_21_ are swapped but the loss function does not fall. So in step 2 the algorithm reverts the array back to the previous state. Then in step 3 *x*_11_ and *x*_31_ are swapped and this time the loss function is lower so the new state is kept.

The overall procedure for simulation of a SNP array is shown in Algorithm 2.

### Production of the GRS from Simulated SNP Arrays

The simulated SNP array is a matrix with values of {0, 1, 2} with each row representing a sample and each column a SNP.

#### Algorithm 2 Simulating SNP arrays

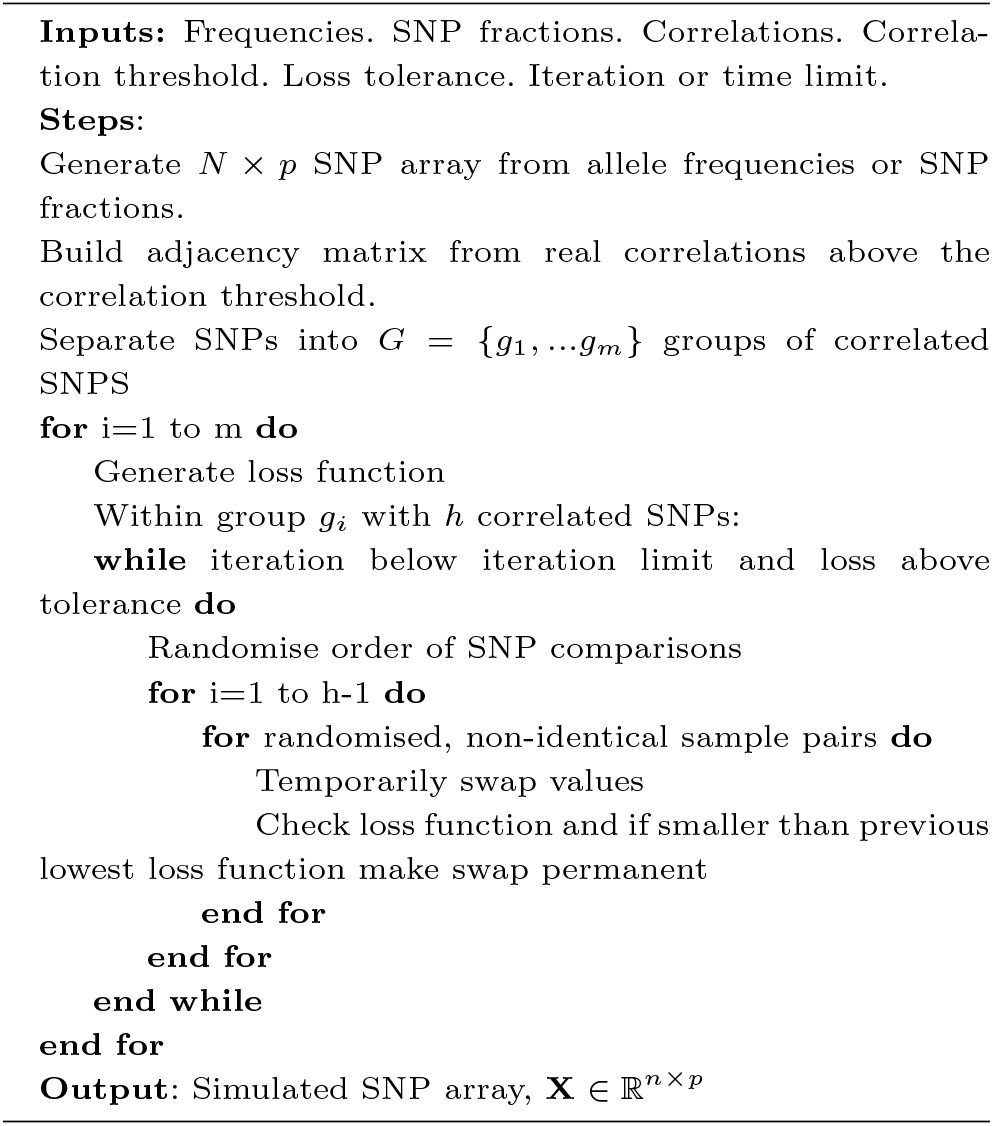

We demonstrate the production of a GRS that is a combination of a linear weighted sum of the alleles with an interaction term between pairs of SNPs. Any GRS of this form, or with just the linear weights could be simulated with this method.

The linear weighted sum requires an effect allele and a weight. The SNP value is switched if the effect allele and the allele representing 2 are not matched. For all p-SNPs the scores are weighted and summed.

For this GRS there is also an interaction term. Here if combinations of pairs (18 in total) of 14 of the SNPs are present together then there is an addition term. Only one interaction term (or none) is permitted per sample and there is a ranking of terms if more than one is present which gives the final interaction term. The interaction term and the linear term are summed together to give the final score.

### Analysis of the Simulated Data

The simulation method produces both a SNP array and a final GRS. To assess how well the approach performs we compare the real data to the simulated data for both 1000G and UKBB datasets. We investigate the final GRS values, relevant sub-parts of the GRS, and comparisons of the SNP arrays themselves.

#### Comparison to final GRS

The 1000G data is a combination of data from different populations around the world. These are combined into 5 super-populations from 26 populations. The simulation method requires the allele frequencies and SNP correlations but these are dependent on which grouping is consider, i.e. whether we use summary statistics from the super-population or population level. We investigate simulations at both levels.

The UKBB data is drawn from the UK so is of people of predominantly Europe descent but there are also substantial numbers of people of other ancestries. We therefore define four different super-populations using PCA and simulate data from those groups. As a fifth group we also use 387 people with T1D from UKBB. This group of European T1D samples (together with the European UKBB super-population) lets us also compare the receiver operating characteristic (ROC) curves and area under the ROC curve (AUC) for the real and simulated data.

#### Comparison to sub-parts of the GRS

We also explore how the model performs at simulating sub-parts of the GRS. Risk of T1D is dominated by the HLA region so we split the GRS into three parts: SNPs from the HLA region, SNPs not from the HLA region, and the interaction term, to assess whether the simulation is correctly estimating each part. In addition, we separately compare the parts of the GRS generated from SNPs that are uncorrelated with any other SNPs in the GRS and those SNPs which are correlated with at least one other SNP in the GRS.

#### Comparison of the SNP arrays

The GRS we consider has 67 SNPs (65 for the 1000G due to missing SNPs). The frequencies of the simulated SNP arrays are defined by the algorithm to match the real frequencies and it (unless it reaches an iteration/time limit before the loss function has fallen below the tolerance limit) adjusts the SNPs with known correlations to other SNPs to be correlated equivalently to the real data (within a tolerance). However, there may be differences between the simulated and real SNP arrays that are not clear from either these summary statistics or the sub-parts of the GRS.

We therefore explore whether the overall structure of the simulated SNP arrays are similar to real SNP arrays by performing principal component analysis (PCA) on the 67 (or 65) SNPs and projecting down onto the first two principal components (PCs). We then investigate similarities and differences between the simulated and real SNP arrays at the level of the first two PCs.

## Results

### Comparison to final GRS

In the left plot of Figure 3 we show the mean simulated GRS against the mean real GRS for the 1000G super-populations and populations. The super-populations are shown with a larger marker. In the right plot we show plots of the mean simulated GRS against mean real GRS for the UKBB super-populations and the T1D population. The real and simulated mean scores for all super-populations and populations are similar.

**Fig. 3.**
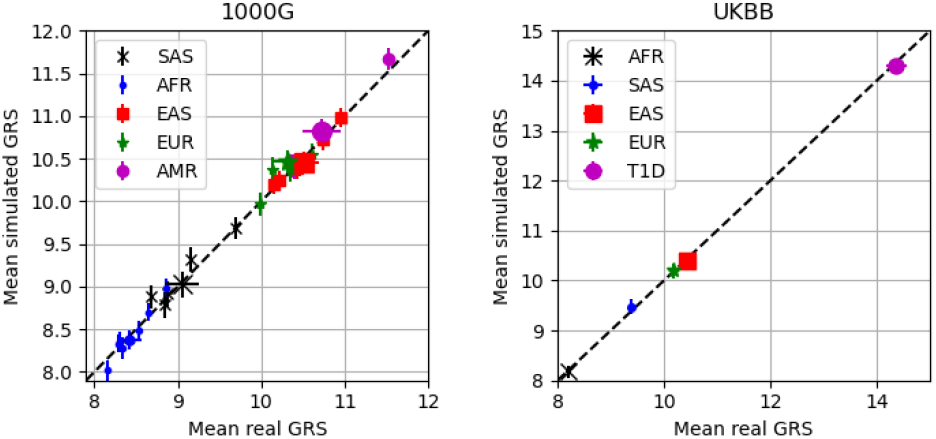
Left) Mean simulated GRS compared to mean real GRS for the 1000G super-populations and populations. The super-populations are marked with a larger marker size. The populations are labelled with their super-population labels, rather that population labels, for clarity. Right) Mean simulated GRS compared to mean real GRS for the UKBB super-populations and the T1D population.

In Figure 4, to the left of the dashed line, we show the distributions of real (left of pair and coloured blue) and simulated (right of pair and coloured orange) GRS for the five populations within the European super-population. Each population has approximately 100 samples within. To the right of the dashed line (violin plots coloured green) we show repeated simulations (labelled S2-S6) of the CEU population (with 99 samples) to demonstrate that there is some variation in the distributions simulated.

**Fig. 4.**
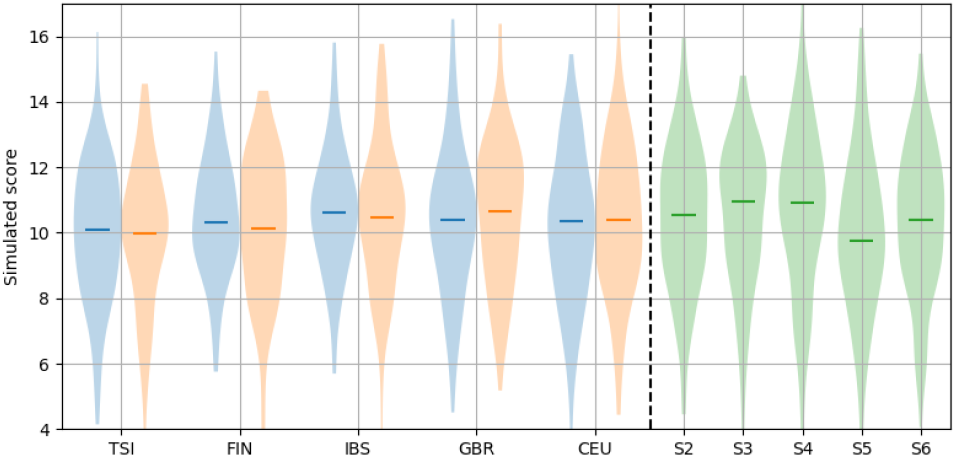
Left of the dashed line are the GRS distributions of the European populations with real GRS (left of pair and in blue) and simulated GRS (right of each pair and in orange) shown side-by-side. TSI (Tuscan from Italy), FIN (Finnish), IBS (Iberians from Spain), GBR (British) and CEU (northern and western European ancestry from Utah, USA) are shown. To the right of the dashed line, and in green, are repeated simulations of the CEU data (labelled S2-S6).

In Table 1 we show the mean and standard deviation of the real (with subscript *R*) and simulated (with subscript *S*) GRSs for the super-populations from 1000G. For the mean and standard deviation (*St. Dev*.) the real and simulated metrics are mostly similar.

**Table 1.**
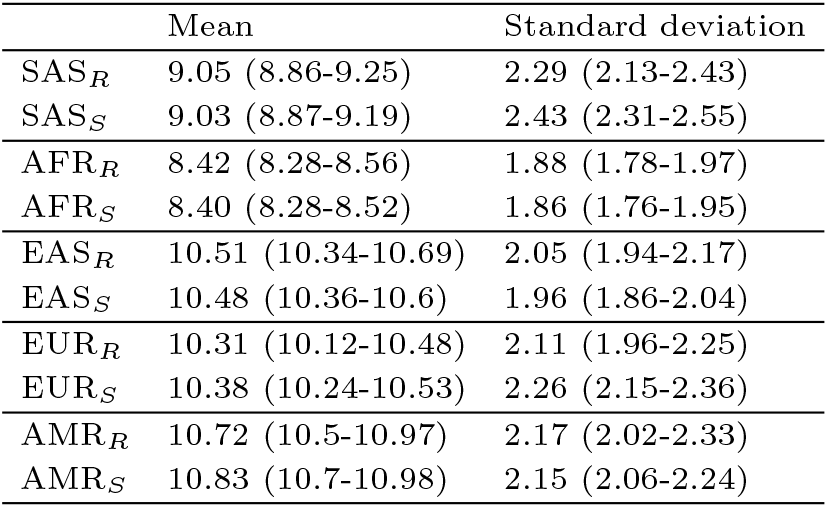
The mean and standard deviation (*St. Dev*.) of the real (denoted by subscript R) and the simulated (denoted by subscript S) GRSs for the 1000G super-populations.

|  | Mean | Standard deviation |
| --- | --- | --- |
| $SAS_R$ | 9.05 (8.86-9.25) | 2.29 (2.13-2.43) |
| $SAS_S$ | 9.03 (8.87-9.19) | 2.43 (2.31-2.55) |
| $AFR_R$ | 8.42 (8.28-8.56) | 1.88 (1.78-1.97) |
| $AFR_S$ | 8.40 (8.28-8.52) | 1.86 (1.76-1.95) |
| $EAS_R$ | 10.51 (10.34-10.69) | 2.05 (1.94-2.17) |
| $EAS_S$ | 10.48 (10.36-10.6) | 1.96 (1.86-2.04) |
| $EUR_R$ | 10.31 (10.12-10.48) | 2.11 (1.96-2.25) |
| $EUR_S$ | 10.38 (10.24-10.53) | 2.26 (2.15-2.36) |
| $AMR_R$ | 10.72 (10.5-10.97) | 2.17 (2.02-2.33) |
| $AMR_S$ | 10.83 (10.7-10.98) | 2.15 (2.06-2.24) |

In the left plot of Figure 5 we show ROC curves and AUC scores for separation of the European T1D and non-T1D data from UKBB for real and simulated data. There are no significant differences between AUC scores for real (denoted in the legend as *R. AUC*) and simulated (denoted in the legend as *S. AUC*) data. In the right plot of Figure 5 we show ROC curves and AUC scores of three simulations of the T1D data (leaving the non-T1D simulated data the same) to show variability in ROC curves from repeated simulations.

**Fig. 5.**
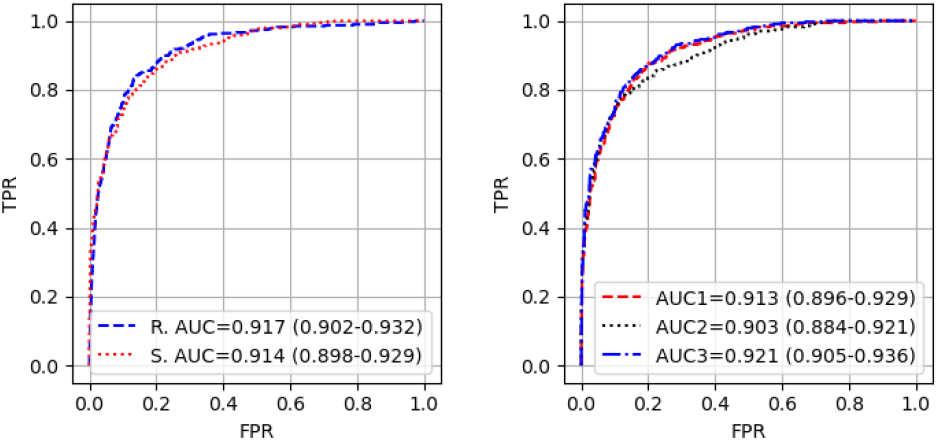
Left) ROC curves for separation of the non-T1D and T1D European populations of UKBB by the real (blue dashed line) and simulated (red dotted line) GRSs. Associated AUC scores are shown for the real (*R. AUC*) and simulated (*S. AUC*) data in the legend. Right) ROC curves for three simulations of the T1D data (with the same simulated non-T1D scores for all three) with AUCs in the legend.

### Comparison to subparts of GRS

The mean sub-parts of the GRS: the linear score of the HLA SNPs (*HLA*), the linear score of the non-HLA SNPs (*Non-HLA*), and the interaction term (*Interaction*) for the 1000G super-populations are shown in Table 2. All sub-parts are equivalent between the simulated and real data with no significant discrepancies.

**Table 2.** Table showing the mean scores for three sub-parts of the GRS on 1000G super-population data. The sub-parts shown are: the linear score of the HLA SNPs (*HLA*), the linear score of the non-HLA SNPs (*Non-HLA*) and the interaction term (*Interaction*). Results are shown for the real scores (subscript *R*) and the simulated scores (subscript *S*).

|  | HLA | Non-HLA | Interaction |
| --- | --- | --- | --- |
| $AFR_R$ | 5.62 (5.45-5.78) | 2.24 (2.19-2.28) | 0.56 (0.47-0.64) |
| $AFR_S$ | 5.62 (5.49-5.75) | 2.23 (2.19-2.26) | 0.55 (0.49-0.61) |
| $SAS_R$ | 6.01 (5.82-6.21) | 3.01 (2.95-3.07) | 0.03 (0.00-0.07) |
| $SAS_S$ | 5.96 (5.82-6.11) | 3.03 (2.98-3.07) | 0.04 (0.01-0.06) |
| $EAS_R$ | 6.98 (6.79-7.18) | 3.40 (3.36-3.45) | 0.12 (0.07-0.18) |
| $EAS_S$ | 6.95 (6.82-7.07) | 3.40 (3.37-3.43) | 0.14 (0.10-0.18) |
| $AMR_R$ | 7.30 (7.04-7.55) | 3.22 (3.14-3.30) | 0.21 (0.11-0.29) |
| $AMR_S$ | 7.34 (7.20-7.47) | 3.24 (3.19-3.28) | 0.22 (0.17-0.27) |
| $EUR_R$ | 6.43 (6.21-6.62) | 3.54 (3.47-3.60) | 0.34 (0.26-0.42) |
| $EUR_S$ | 6.45 (6.30-6.61) | 3.52 (3.47-3.57) | 0.32 (0.27-0.38) |

In Table 3 we show the number of SNPs which are correlated with at least one other SNP in the GRS (*Correlated number*) and the mean summed scores for both the SNPs which are correlated with at least one other SNP and SNPs which are not correlated with any other SNPs in the GRS for the 1000G super-populations. The simulated GRS produces similar mean scores to the real data for both the correlated and non-correlated SNPs.

**Table 3.**
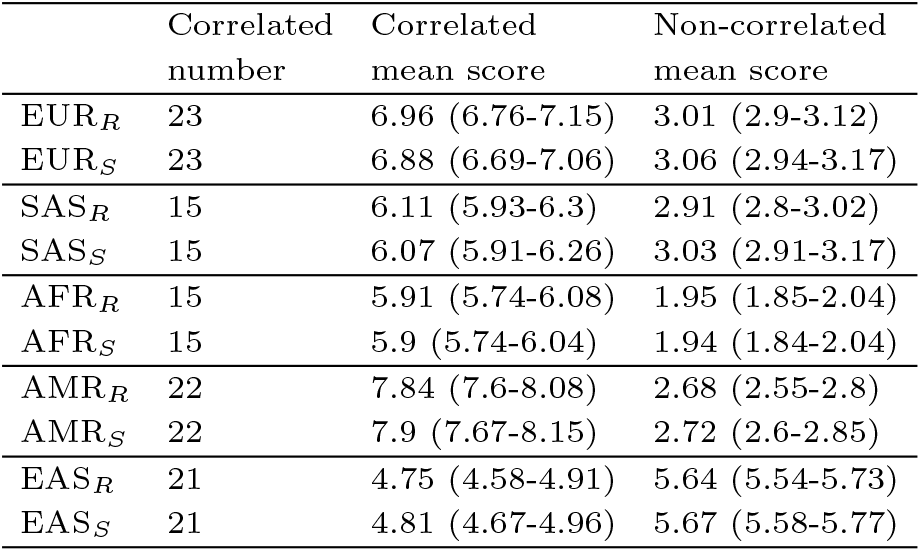
Mean scores for the real (subscript *R*) and simulated (subscript *S*) 1000G super-population data when separating SNPs into those that are correlated (*Correlated mean score*) and non-correlated (*Non-correlated mean score*). The number of SNPs that are correlated with at least one other SNP in the GRS are also shown (*Correlated number*).

|  | Correlated number | Correlated mean score | Non-correlated mean score |
| --- | --- | --- | --- |
| EUR<sub>R</sub> | 23 | 6.96 (6.76-7.15) | 3.01 (2.9-3.12) |
| EUR<sub>S</sub> | 23 | 6.88 (6.69-7.06) | 3.06 (2.94-3.17) |
| SAS<sub>R</sub> | 15 | 6.11 (5.93-6.3) | 2.91 (2.8-3.02) |
| SAS<sub>S</sub> | 15 | 6.07 (5.91-6.26) | 3.03 (2.91-3.17) |
| AFR<sub>R</sub> | 15 | 5.91 (5.74-6.08) | 1.95 (1.85-2.04) |
| AFR<sub>S</sub> | 15 | 5.9 (5.74-6.04) | 1.94 (1.84-2.04) |
| AMR<sub>R</sub> | 22 | 7.84 (7.6-8.08) | 2.68 (2.55-2.8) |
| AMR<sub>S</sub> | 22 | 7.9 (7.67-8.15) | 2.72 (2.6-2.85) |
| EAS<sub>R</sub> | 21 | 4.75 (4.58-4.91) | 5.64 (5.54-5.73) |
| EAS<sub>S</sub> | 21 | 4.81 (4.67-4.96) | 5.67 (5.58-5.77) |

### Comparison to SNP arrays

In Figure 6 we show a comparison of the real (left plot) and simulated (right plot) 1000G data when applying PCA to the whole dataset (2503 samples) and the 65 SNPs. The first two principal components are shown and the colours represent the different super-populations. The pattern of the real and simulated data is similar with comparable separation of the super-populations.

**Fig. 6.**
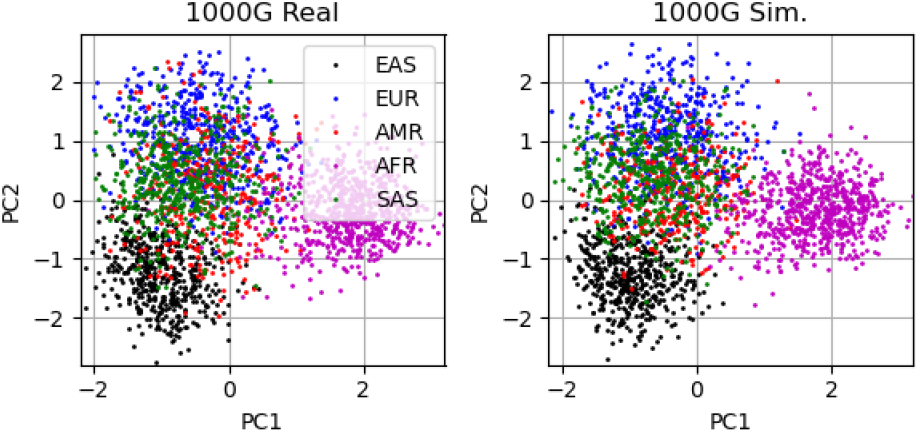
PCA applied to the real (left) and simulated (right) whole 1000G dataset (2503 samples). The first two principal components are shown and the colours represent the different super-populations.

In Figure 7 we demonstrate, using the AMR super-population, the importance of which summary statistics are used to simulate the SNP arrays. The top left plot shows the first two principal components applied to the 1000G AMR real data while the top right is the simulated SNP array when the simulations were generated using summary statistics at the super-population level. The bottom left shows simulated PCA of the SNP array when the four different populations making up the super-population are generated separately (with summary statistics at the population level) and concatenated together before PCA is applied. The bottom right plot shows the same plot as the bottom left but with markers and colours representing each population. The data is more spread out when the summary statistics are used from the populations rather than the super-populations.

**Fig. 7.**
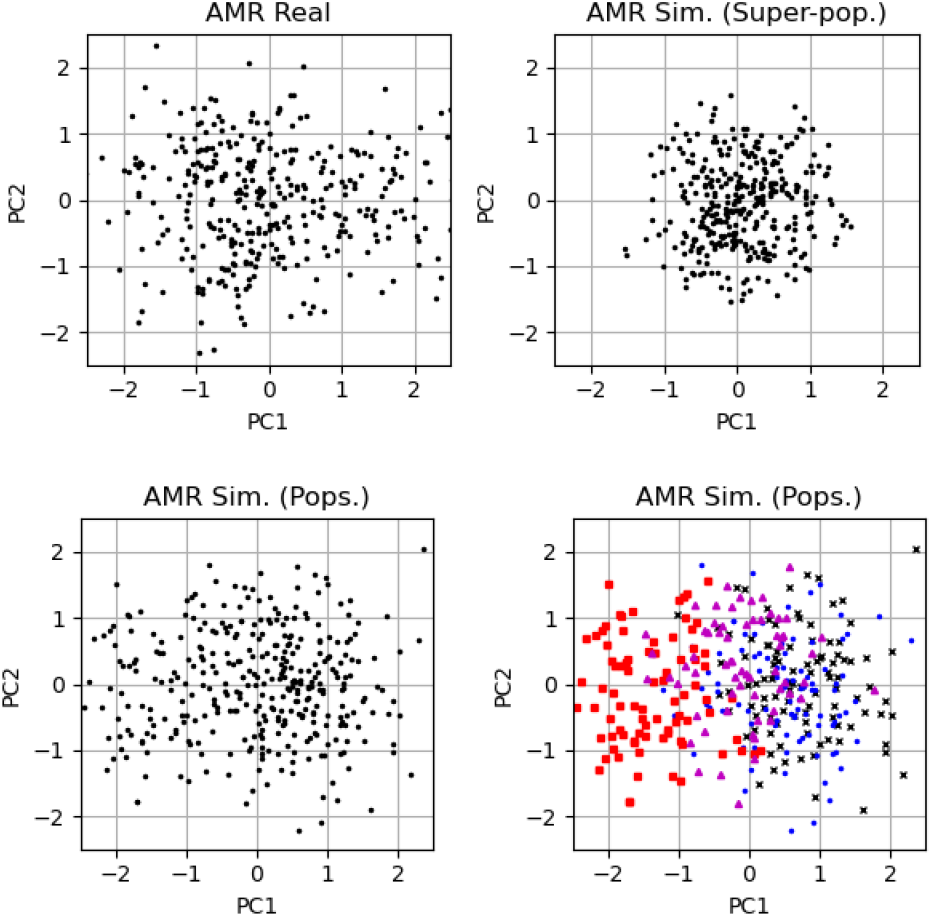
The first two principal components of the real and simulated 65-SNP T1D GRS for the AMR super-population. Top left plot is the real SNP PCA. Top right is the simulated data when generated from the summary statistics on the super-population as a whole (labelled *superpop*.). The bottom left is simulated data when four separate SNP arrays are generated for the four populations making up the super-population and then the SNP arrays are concatenated together before PCA is applied (labelled *pops*.). The bottom right plot shows the same as the bottom left but with the populations denoted with different marker shapes and colours.

In Figure 8 we show that the simulation model can produce structure in the first two principal components similar to the real data. The top left and top right plots show the first two principal components of the real and simulated data of the AFR super-population of the 1000G data with the seven populations represented by different marker colours and shapes. In the bottom row of Figure 8 we show a partial cause of the structure in the AFR super-population first two principal component plot. The left and right plots are the first two principal components of the real data and the simulated data respectively. The three marker colours/shapes represent those samples which have one SNP (rs17843689) either as 0, 1, 2 (on the effect allele).

**Fig. 8.**
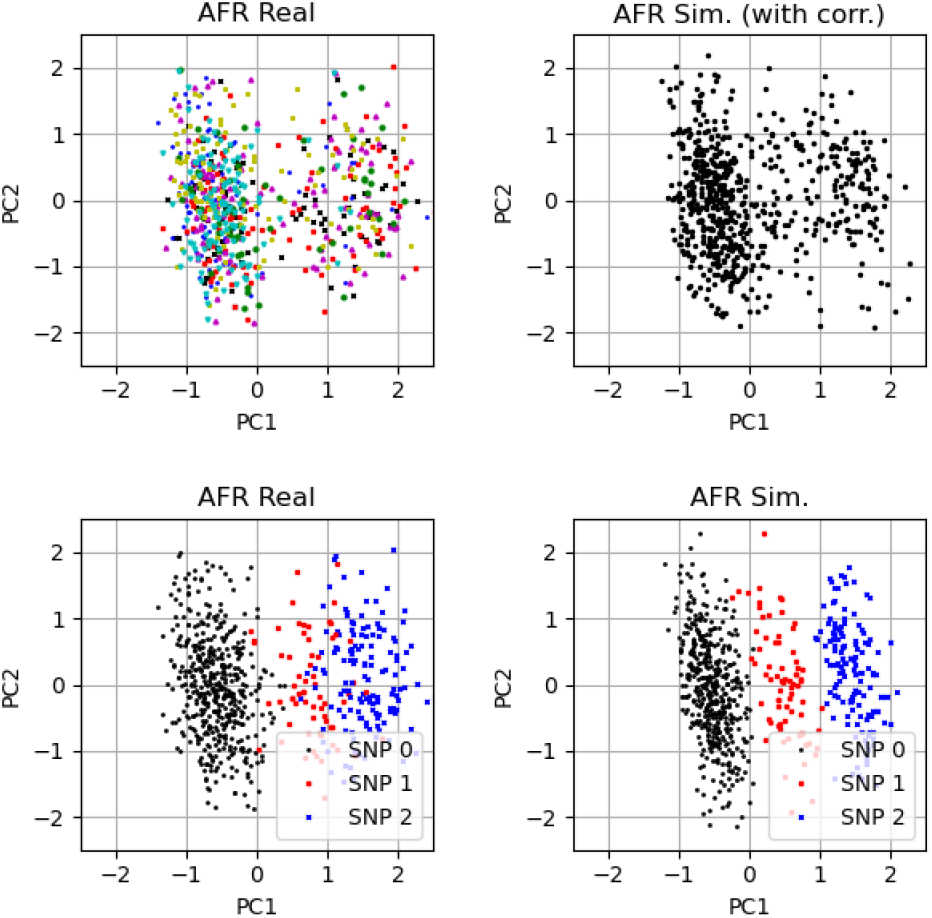
The top left and right plots are the first two principal components of the AFR super-population data from the 1000G for real and simulated data respectively. Similar structure is visible in both PCA plots. The different colours of the markers are different populations. The bottom left (real data) and right (simulated data) plots show the relationship between a sample from the AFR super-population of the 1000G having a SNP (rs17843689) variant and its location in PCA space. The colours represent whether the sample has a 0, 1, 2 value for that SNP, labelled as SNP 0, SNP 1, SNP 2 respectively.

In Figure 9 we show the first two principal components for the European T1D and non-T1D data from the UKBB dataset. The red crosses are the T1D data and the blue dots are 1000 random samples from the non-T1D data. The left and right plots are the real and simulated data respectively. For both the real and simulated data the T1D samples occupy similar parts of the first two principal components space.

**Fig. 9.**
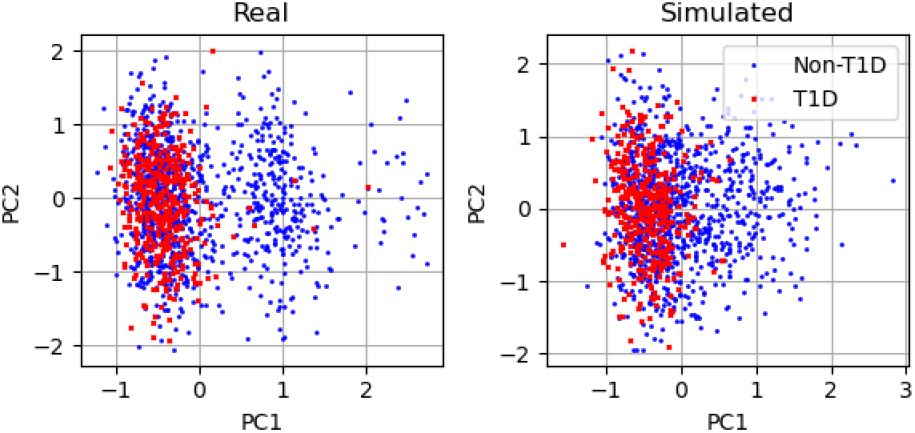
The first two principal components for the European T1D and non-T1D data from the UKBB dataset. The red crosses are the T1D data and the blue dots are 1000 random samples from the non-T1D data. Left) Real data. Right) Simulated data.

## Discussion

The overall aim of this work was to simulate GRSs without access to the original genotype data. The quality of the simulated data was compared to real data both via the summary statistics relating to the GRS and the SNP arrays.

The mean, median and standard deviation of the simulated GRSs are similar to the equivalents for the real data for both the 1000G data and UKBB. In addition, an AUC produced by considering T1D cases in UKBB and comparing to non-T1D controls are similar for simulated and real data. Therefore, summary statistics from the simulated data would produce the similar conclusions as using real genetic data. Given a common use of GRSs is to calculate discrimination between cases and controls via AUC, we have demonstrated that the simulated data would produce similar AUC as the real data.

From the summary statistics produced by the simulated data it could be concluded that the simulation approach produces a GRS similar to that produced on real data. However, it is important to understand if there is variation that is hidden when considering summary statistics, which might result in false final results in other circumstances.

To investigate the simulation quality further we considered both sub-parts of the GRS and the generated SNP arrays themselves. For the sub-parts of the GRS we looked at the generation of the HLA, non-HLA and interaction parts. There was no difference in the mean scores for these three subparts of the GRS between simulated and real data for either the 1000G data or the UKBB data.

We also considered the SNPs that are correlated and the non-correlated SNPs. If the models were not correctly producing the correlations between the SNPs then the differences might be expected to occur in those SNPs which were correlated with at least one other SNP as these needed to be optimised by the algorithm. However, we see no differences between simulated and real mean scores generated using either correlated or non-correlated SNPs.

The metrics summarising the GRS, even sub-parts of the GRS, might conceal variations in the distributions of the GRSs. These distributions can vary between different runs of the simulation model (see Figures 4) making it difficult to be confident that the simulated models are producing the same distributions as the real data. None of the variation in distributions is likely to have an effect on any conclusions drawn though and the real data is also drawn from a much larger real distribution, but we cannot be certain that our simulated model is correctly estimating the entire distribution.

When considering the relationship between real and simulated data at the SNP-level, the frequencies of the SNPs are defined to be correct and the correlations are optimised by the algorithm. The pattern of SNP values overall might be different though. To investigate these patterns between the SNPs we took the first two principal components of the real and simulated SNP arrays.

When performing PCA for all the 1000G data and comparing the real and simulated first two principal components (see Figure 6) the patterns produced are similar for the real and simulated SNP arrays. We see the same pattern of separation of super-populations which means that the simulation is capturing similar distributions of SNPs at the super-population level.

We demonstrated that the level of the population of the SNP summary statistics is particularly important for some of the datasets. In particular for varied populations like the 1000G AMR super-population, where the simulated SNP arrays are insufficiently separated if we take frequencies and correlations at the super-population level (see Figure 7). By simulating the SNP arrays using the summary statistics at the population level we produce distributions across the first two principal components that are more similar to the real SNP arrays.

The simulation model also generates some structures in PCA space which are similar to real data. We showed this in Figure 8 where the PCA plots of real and simulated data have some structural similarities. A partial cause of this structure was also shown in Figure 8 where the value of one SNP was driving a lot of the variance in the first principal component.

We also simulated the T1D data at the SNP-array level where the T1D sits in the same part of the PCA space as the real T1D data. There is some variation in the structure in the first two PCs here.

Overall, the simulated GRS method produces data which appears to be similar to the real data. This is the case both if we consider summary statistics (mean, median and standard deviation) and discrimination power of the GRS (AUC between T1D cases and non-T1D controls). We also see no differences in sub-parts of the GRS. At the SNP-array level (considered as the first two principal components) we find similar distributions between real and simulated data.

While we have demonstrated that the model produces simulated data that is similar to the real data we cannot be sure that there are not failure cases, for some reason where the simulation method would not work. These is some variation in the principal components of the SNP arrays which may be statistical variation or might be real differences in structure. There are also some differences in the summary statistics, which could be for statistical sampling reasons or may be due to genuine variation between real and simulated data.

Another issue, is in the level of population required - we demonstrated that the super-population of the Americans from 1000G is not the correct level to draw summary statistics from. There may be subclasses below that which would be more appropriate.

If there are too many correlated SNPs, or if there are correlated SNPs that have low frequencies, the algorithm can struggle to find an array that fully matches the correlated SNPs. There can also be spurious correlations that can be imposed by the algorithm. These effects are not significant in the datasets we tested the algorithm on.

## Conclusions

Our approach allows the generation of simulated SNP arrays for GRS generation as well as the final GRS. The simulated scores show either the same or highly similar structure to real scores generated from genotyped or imputed data. Our method allows comparisons to be made by the simulation of GRSs from summary statistics without the need for access to genotyped arrays.

## Data Availability

The study used only openly available human data. The three datasets used were: 1000G available at https://www.internationalgenome.org/; UK Biobank available at https://www.ukbiobank.ac.uk/; T1DGC available at https://repository.niddk.nih.gov/studies/t1dgc/.

https://repository.niddk.nih.gov/studies/t1dgc/

## Acknowledgements

This study was supported by a grant from the Randox Corporation.

This study was supported by the National Institute for Health and Care Research Exeter Biomedical Research Centre. The views expressed are those of the author(s) and not necessarily those of the NIHR or the Department of Health and Social Care.

## Notes

### Competing Interest Statement

The authors have declared no competing interest.

